# Time to negative PCR from symptom onset in COVID-19 patients on Hydroxychloroquine and Azithromycin - A real-world experience

**DOI:** 10.1101/2020.08.05.20151027

**Authors:** Sarfraz Saleemi, Abdulrahman Alrajhi, Mohammed Alhajji, Areej Alfattani, Faisal Albaiz

**Author notes:** Corresponding author: ^1^Sarfraz Saleemi, Section of Pulmonary Medicine, Department of Medicine MBC 46, King Faisal Specialist Hospital & Research Center, Riyadh 11211, PO Box 3354, Saudi Arabia.

## Abstract

**Background:** The role of hydroxychloroquine (HCQ) and azithromycin in the treatment of COVID-19 and its effect on SARS-CoV-2 viral clearance is not known.

**Methods:** This is a retrospective observational study to assess the effect of HCQ and Azithromycin on duration from symptom onset to negative SARS-CoV-2 PCR using nasopharyngeal swab in hospitalized patient with COVID-19. Eighty-five patients were included in the study, 65 in HCQ (Hydroxychloroquine + Azithromycin) and 20 in non-HCQ group. Measurement of duration from symptom onset to negative PCR and effect of gender, age and disease severity on time to viral clearance was measured.

**Results:** Median time to negative PCR in HCQ group was 23 days (IQR: 9, Mean 24+8, N=65) compared with non-HCQ group, 19 days (IQR: 8, Mean 18±6, N=20), *(p* <0.05). Forty-one (63%) patients in HCQ group and all patients (100%) in non-HCQ group had mild disease. Multivariate regression model (F=6.8, *P*<0.002, R^2^=0.20) shows that being in HCQ group would delay the time to negative PCR by 7 days (95%CI: 2-12) and with every year increase in the age, the time to negative PCR would be delayed by 0.12 days (95%CI: 0.017-0.22). Among HCQ sub-groups, gender and disease severity had no effect on duration (*p* 0.142 and 0.156 respectively) but older patients ≥60 year had longer duration compared to patients <60 year of age although *p* value did not reach significance (*p* 0.073). Median time to negative PCR in mild- HCQ group (23 days, IQR: 9, Mean 23+8, N=41) was longer when compared with non-HCQ group *(p* <0.05). On day 28, all patients in non-HCQ group had negative PCR while only 50/65 (77%) were negative in HCQ group.

**Conclusion:** Hydroxychloroquine (HCQ) and azithromycin delay SARS-CoV-2 virus clearance in hospitalized patients with COVID-19 and it is correlated with older age. Larger studies are needed to confirm this finding.

## Introduction

COVID-19 caused by SARS CoV-2 is a highly contagious disease that emerged in the late 2019 in Wuhan, China.^1,2^ The incubation period for COVID-19 is within 14 days with the median incubation period 4 days (range 2-7 days). The vast majority of cases will make a full recovery, though this may take several weeks, particularly in severe cases. In a minority of cases, COVID-19 has been associated with rapid progression to acute respiratory distress syndrome (ARDS), multiple organ failure and death. The data from initial outbreak in China reported 13.8% of cases with severe disease, and 6.1% with critical course.^3^ The case fatality ratio is currently unknown, but it is estimated to be within the range of 0.5-4%. The discharge criteria after hospital admission varies from region to region and has been changing with time as virus kinetics are better understood. A study of nine patients with COVID-19 shows that virus obtained from patients after eight days of illness did not grow in culture or yield sub-genomic mRNA which is only present when a virus is replicating. This study indicates that even though the viral RNA is detected by real time reverse transcription polymerase chain reaction (RT-PCR), it may not be active or infectious after eight days of illness.^4^ Current recommendations by World Health Organization (WHO) for discharging patients from isolation regardless of location or disease severity are 10 days after symptom onset, plus 3 additional days without symptoms.^5^ Several health institutions still use SARS-CoV-2 RT-PCR based criteria for discharge.

Hydroxychloroquine (HCQ), a widely used anti-malarial drug gained popularity in the treatment of COVID-19 in the beginning of the pandemic due to its immunomodulatory properties and its inhibitory effect on SARS COV-2 in vitro.^6^ Based on pharmacokinetic models, a dose of 400 mg twice a day followed by 200 mg twice a day was recommended.^7^ Time to negative RT-PCR for SARS COV-2 is variable and may be affected by the current experimental medications by reducing viral load and viral clearance. An open labeled French study by Gautret et al. showed that hydroxychloroquine treatment is significantly associated with viral load reduction/disappearance in COVID-19 patients and its effect is reinforced by azithromycin. At day 6 post-inclusion, 70% of hydroxychloroquine-treated patients were virologically cured comparing with 12.5% in the control group (*p*= 0.001).^8^ A small Chinese randomized controlled trial of 30 treatment naïve patients showed that on day 7, nucleic acid of throat swab was negative in 13 (86.7%) cases in HCQ group and 14 (93.3%) cases in the control group (*p* >0.05).^9^ Another retrospective study of 301 COVID-19 patients from China showed that the median period from symptoms onset to negative SARS-CoV-2 RT-PCR was 20 days (IQR, 17-24; N = 216). Older patients ≥65 year had longer duration to negative RT-PCR compared to younger patients (22 days vs 19 days, *p* 0.015).^10^

This retrospective study was conducted to assess the duration from symptom onset to negative RT-PCR in patients with COVD-19 who were treated with Hydroxychloroquine and Azithromycin (HCQ group) compared to those who did not receive the treatment (non-HCQ group).

## Methods

### Study design

This is a retrospective study of laboratory confirmed COVID-19 patients who were admitted to a tertiary care facility. The diagnosis was confirmed by Real Time Reverse Transcription Polymerase Chain Reaction (RT-PCR) for SARS-CoV2 by taking nasopharyngeal swab and using PCR testing kits by Altona (Hamburg-Germany), approved by government health regulatory authorities. Adult patients who had negative RT-PCR at the time of discharge were included in the study. Pediatric patients <14 year of age and those who were discharged to home isolation without being admitted to hospital were excluded from the study.

### Data collection

The data was extracted from electronic medical record and REDCap (Research Electronic Data Capture) platform, which is password protected and regulated by Research Advisory Council of the institution and confidentiality is insured by restricting access to only approved project investigators. Data analyzers other than approved investigators did not have access to medical record numbers of the study subjects. Data was collected about demographics, clinical, laboratory, SARS-CoV2 PCR, therapeutic agents, symptom onset, admission and discharge dates. Severity of disease was classified into four groups as per institution guidelines. Stage A - patient with no relative clinical signs or symptoms but positive SARS-CoV2 PCR, Stage B - patient with upper respiratory tract infection symptoms and/or other symptoms including fever and gastrointestinal symptoms without evidence of pneumonia, Stage C - patient with radiology evidence of lower respiratory tract infection and/or hypoxemia with oxygen saturation ≤93% at rest breathing room air or drop in oxygenation compared to baseline but not requiring intensive care unit (ICU) and Stage D was categorized as patients requiring ICU admission. In this study, stage A and B are categorized as mild and stage C and D as severe disease. The study subjects were grouped into Hydroxychloroquine (HCQ) and non-HCQ. Patients who received combination of Hydroxychloroquine and Azithromycin or Hydroxychloroquine alone were included in HCQ group and those who were not prescribed Hydroxychloroquine were included in non-HCQ group. The duration from symptom onset to negative PCR was determined by the first negative RT-PCR if only one PCR was done before discharge and in the event, two consecutive RT-PCR were done 24-hour apart, the 2^nd^ PCR was used as a reference point. Some patients who had positive result after first negative result were tested again and the repeat negative RT-PCR was considered for calculation of duration. Dynamic profile of SARS-CoV2 PCR in both groups and impact of disease severity, gender and age on duration to negative PCR was documented.

### Statistical analysis

Data was summarized by using descriptive statistics. Results were reported as median with inter-quartile range and mean with standard deviation and categorical variable were calculated as counts (n) and percentages (%). Difference between groups were analyzed using Mann-Whitney U test, T-test and two tailed Z-test where appropriate. Multivariate linear regression (stepwise approach) for the time to PCR negative was applied to assess the effect of the significant co-factors in a prediction model. All *p* values were reported with significance level of 0.05.

### Ethical consideration

The study was granted approval by the institutional review board and written informed consent was waived in view of emerging pandemic and retrospective noninterventional study.

## Results

### Demographic and clinical characteristics

Eighty-five patients were included in the study, 65 in HCQ and 20 in non-HCQ group. Mean age in HCQ group was 48 ±18 years and 27 (42%) were males while 46 ± 11 years and 13 (65%) in non-HCQ group respectively. Forty-three (66%) patients had co-morbid medical conditions in HCQ group vs 8 (40%) in non-HCQ group (*p*<0.05). HCQ group had 41(63%) patients with mild and 24 (37%) severe disease while none of the patients in non-HCQ group had severe disease. Fever, cough, sore throat and fatigue were common in HCQ vs non-HCQ group (*p* <0.05). C-reactive protein and D-dimer levels were higher in HCQ group (*p* <0.05) and there was no difference in Neutrophil/Lymphocyte ratio and Ferritin level between the groups. Sixty-three (97%) patients received combination of Hydroxychloroquine and Azithromycin in HCQ group. Hydroxychloroquine was prescribed at a dose of 400 mg twice a day for first day followed by 200 mg twice a day for four days. In addition, 28 (43%) received antibiotics, 10 (15%) Lopinavir/Ritonavir(Kaletra), 7 (11%) Ribavirin, 5(8%) Tocilizumab, and 1 (2%) Intravenous Immunoglobulin (IVIG). In non-HCQ group, 2 (10%) patients received Azithromycin, 1 (5%) each antibiotic, Lopinavir/Ritonavir(Kaletra) and Interferon.(Table 1) HCQ-mild and non-HCQ groups were comparable except that the symptoms of fever, cough, sore throat and fatigue were more common and C-reactive protein and D-dimer levels were higher in HCQ-mild group (*p* <0.05) (Table 2)

**Table 1.**
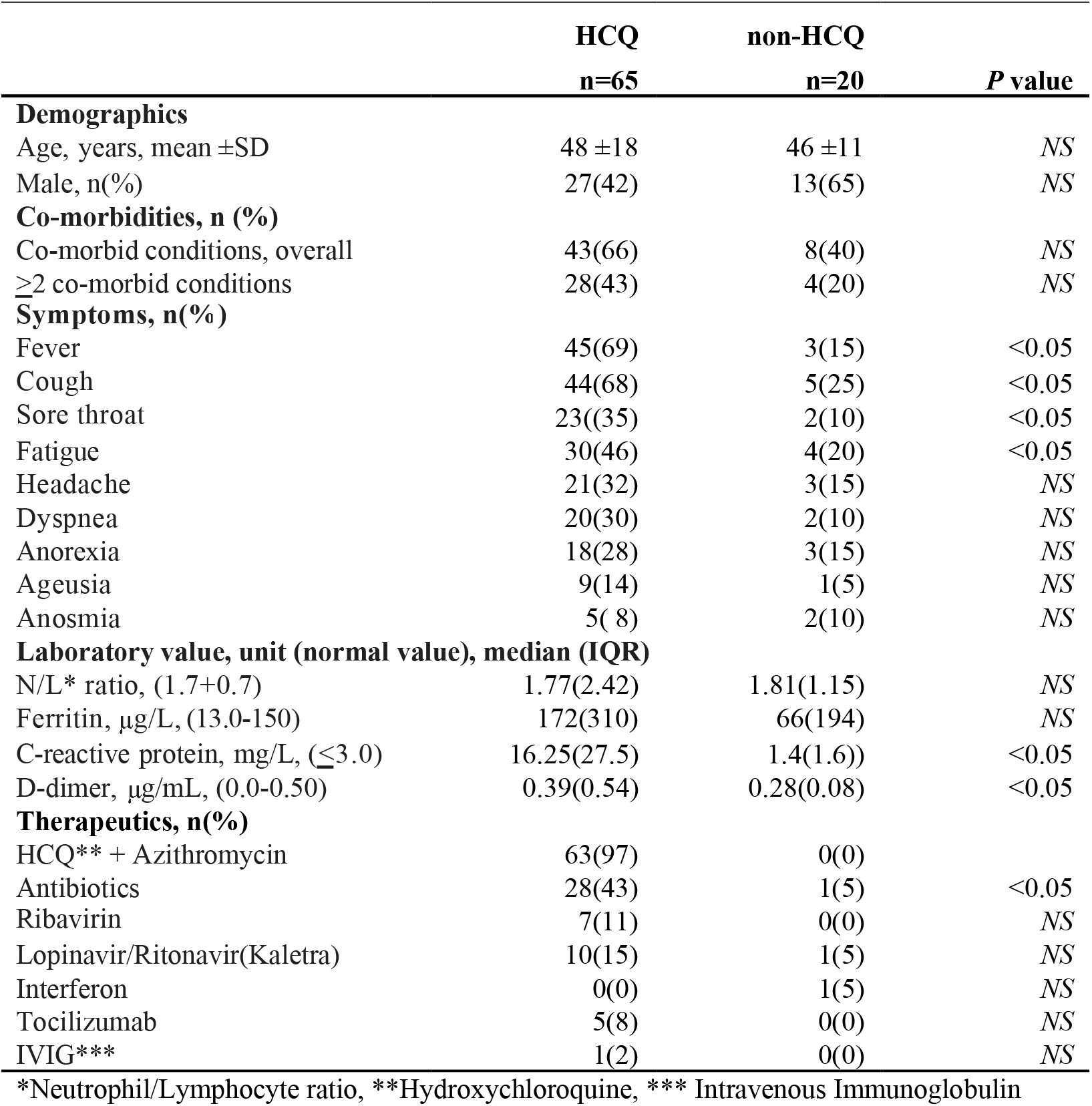
Demographic and clinical characteristics, HCQ and non-HCQ group

|  | HCQ<br>n=65 | non-HCQ<br>n=20 | P value |
| --- | --- | --- | --- |
| **Demographics** |  |  |  |
| Age, years, mean ±SD | 48 ±18 | 46 ±11 | NS |
| Male, n(%) | 27(42) | 13(65) | NS |
| **Co-morbidities, n (%)** |  |  |  |
| Co-morbid conditions, overall | 43(66) | 8(40) | NS |
| $\geq 2$ co-morbid conditions | 28(43) | 4(20) | NS |
| **Symptoms, n(%)** |  |  |  |
| Fever | 45(69) | 3(15) | <0.05 |
| Cough | 44(68) | 5(25) | <0.05 |
| Sore throat | 23(35) | 2(10) | <0.05 |
| Fatigue | 30(46) | 4(20) | <0.05 |
| Headache | 21(32) | 3(15) | NS |
| Dyspnea | 20(30) | 2(10) | NS |
| Anorexia | 18(28) | 3(15) | NS |
| Ageusia | 9(14) | 1(5) | NS |
| Anosmia | 5( 8) | 2(10) | NS |
| **Laboratory value, unit (normal value), median (IQR)** |  |  |  |
| N/L* ratio, (1.7+0.7) | 1.77(2.42) | 1.81(1.15) | NS |
| Ferritin, μg/L, (13.0-150) | 172(310) | 66(194) | NS |
| C-reactive protein, mg/L, ($\leq 3.0$) | 16.25(27.5) | 1.4(1.6) | <0.05 |
| D-dimer, μg/mL, (0.0-0.50) | 0.39(0.54) | 0.28(0.08) | <0.05 |
| **Therapeutics, n(%)** |  |  |  |
| HCQ** + Azithromycin | 63(97) | 0(0) |  |
| Antibiotics | 28(43) | 1(5) | <0.05 |
| Ribavirin | 7(11) | 0(0) | NS |
| Lopinavir/Ritonavir(Kaletra) | 10(15) | 1(5) | NS |
| Interferon | 0(0) | 1(5) | NS |
| Tocilizumab | 5(8) | 0(0) | NS |
| IVIg*** | 1(2) | 0(0) | NS |

\*Neutrophil/Lymphocyte ratio, \*\*Hydroxychloroquine, \*\*\* Intravenous Immunoglobulin

**Table 2.**
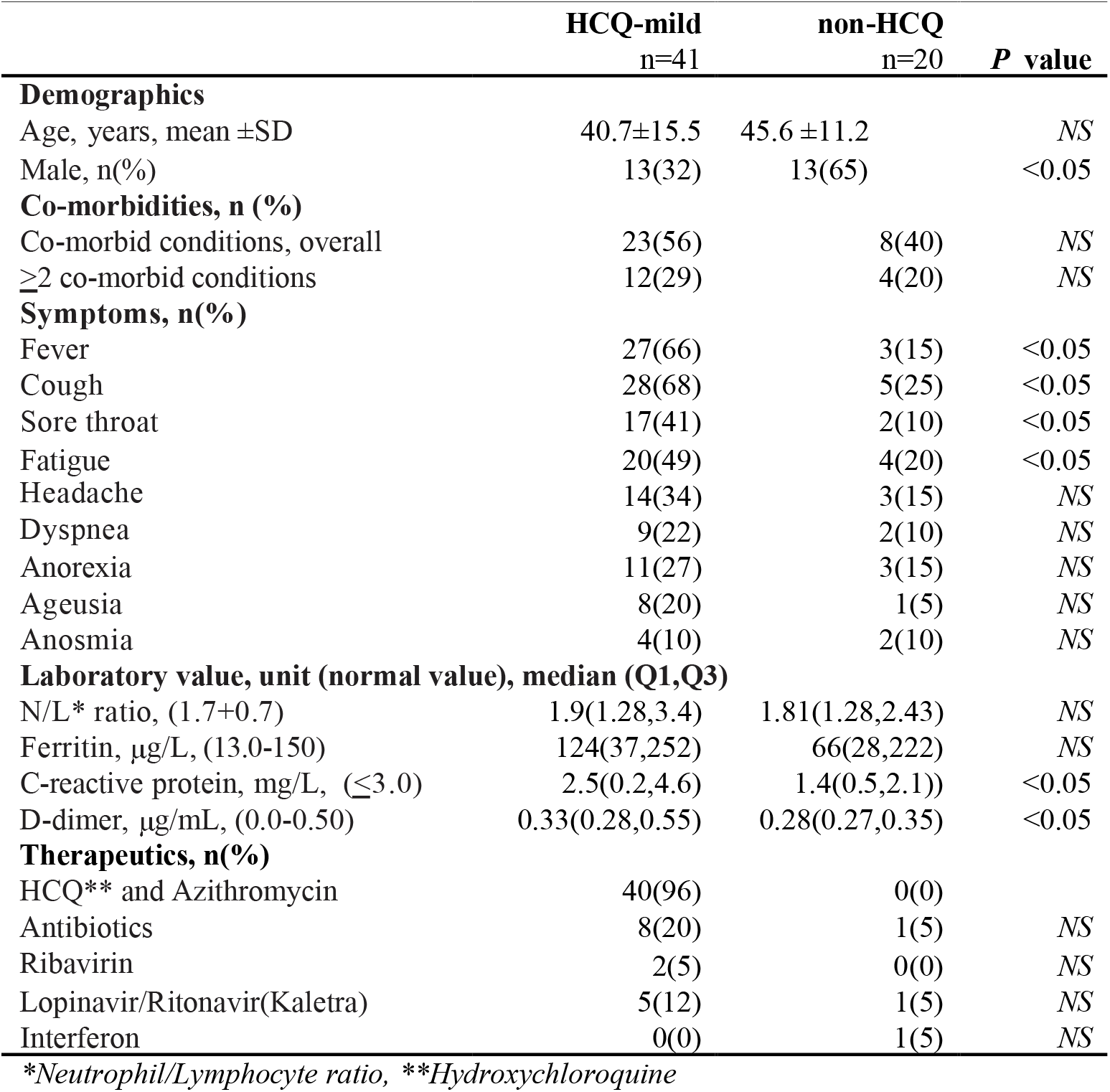
Demographic and clinical characteristics of HCQ-mild and non-HCQ

### Dynamics of SARS-CoV2 PCR

Median duration from symptom onset to negative PCR was 23 days (IQR:9) in HCQ group and 19 days (IQ:8) in non-HCQ group (*p* <0.05). (Fig 1) On day 28, all patients were negative for SARS-CoV2 PCR in non-HCQ group while only 50/65 (77%) in HCQ group (*p* <0.05). (Table 3) The positive rate of PCR at day 0-7 was 100% in HCQ group vs 90% in non-HCQ group, day 8-14, 90% vs 70%, day 15-21, 63% vs 25% and at day 22-28, 23% vs 0% respectively. (Fig 2)

**Table 3.** Time to negative PCR from symptom onset

|  | HCQ<br>Overall<br>n=65 | HCQ<br>Mild<br>n=41 | HCQ<br>Severe<br>n=24 | HCQ<br><60yr<br>n=48 | HCQ<br>≥60yr<br>n=17 | non-HCQ<br>n=20 |
| --- | --- | --- | --- | --- | --- | --- |
| **Median(IQR),days** | 23(9) | 23(9) | 25(9) | 23(10) | 25(8) | 19(8) |
| **Negative rate of SARS-CoV-2 RT-PCR, n(%)** |  |  |  |  |  |  |
| Day 0-7 | 0(0) | 0(0) | 0(0) | 0(0) | 0(0) | 2(10) |
| Day 8-14 | 6(9) | 2(8) | 4(10) | 6(16) | 0(0) | 6(30) |
| Day 15-21 | 24(37) | 8(33) | 16(39) | 19(36) | 5(29) | 15(75) |
| Day 22-28 | 50(77) | 18(75) | 32(78) | 40(83) | 11(67) | 20(100) |
| Day 29-35 | 58(89) | 20(83) | 38(93) | 45(94) | 13(76) |  |
| Day 36-42 | 64(98) | 24(100) | 40(98) | 48(100) | 16(94) |  |
| Day 43-49 | 65(100) |  | 41(100) |  | 17(100) |  |

**Fig 1.**
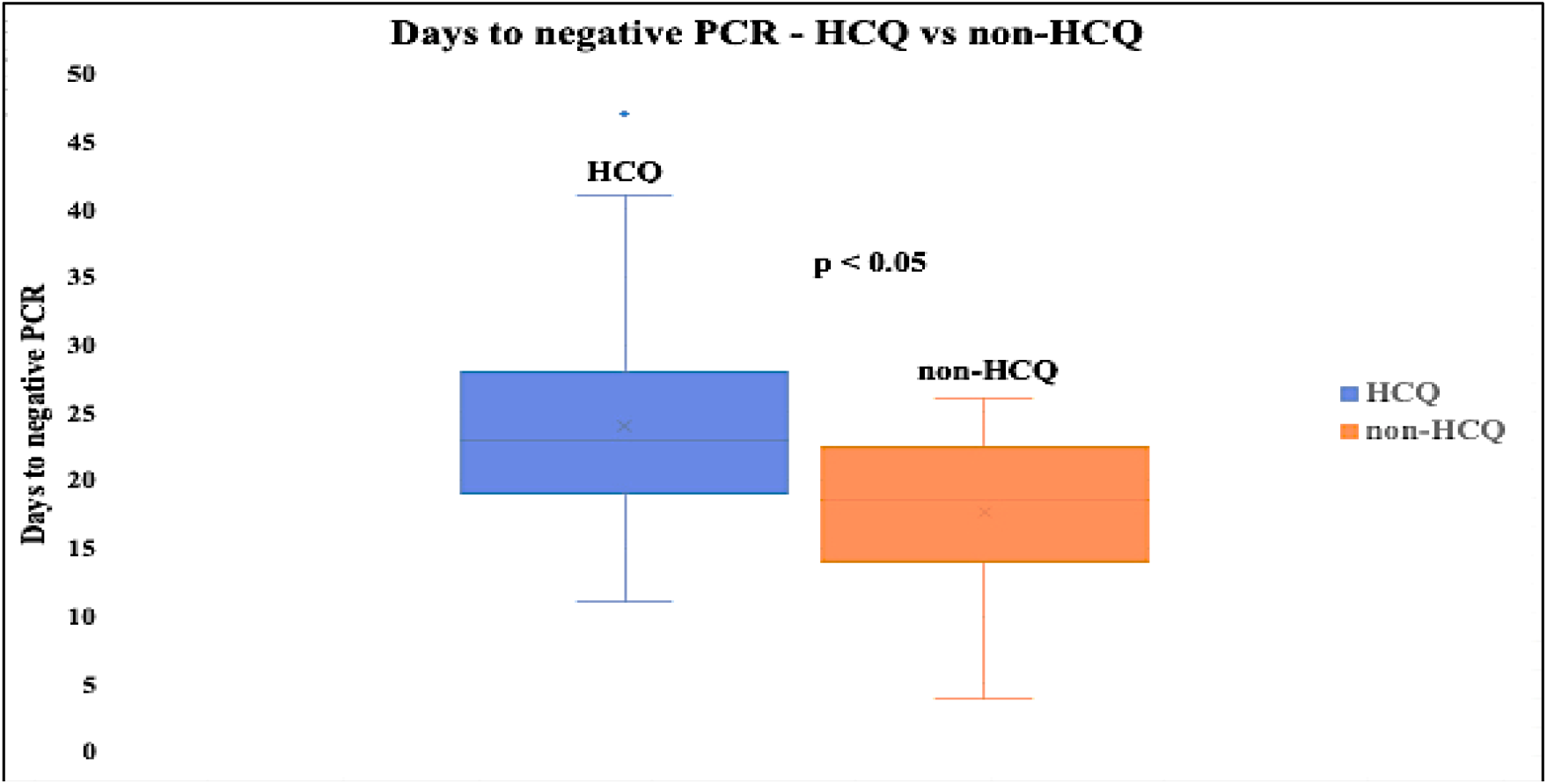
Time to negative PCR from symptom onset.

**Fig 2.**
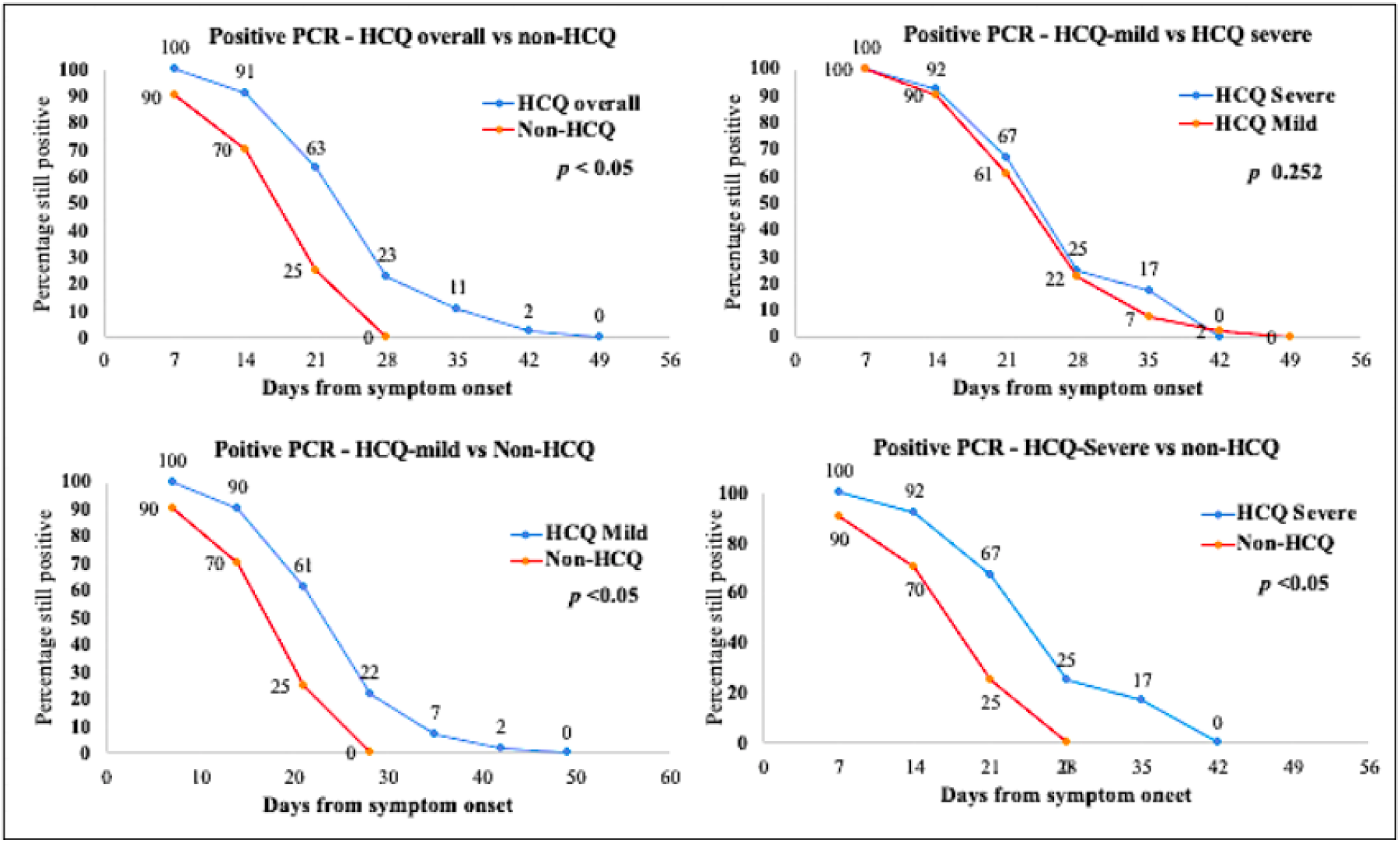
Positive PCR rate.

### Effect of disease severity and demographics on virus dynamics

Among HCQ sub-groups, there was no significant difference in time to negative PCR between mild and severe disease, males and females, but older patients ≥60 year had longer median duration to negative PCR compared to younger patients <60 years although *p* value did not reach significance *(p* 0.073). The median time to negative PCR was 25 days (IQR:9.2) for the severe vs 23 days (IQR:9) for the mild HCQ cases. (*p =NS)*. in addition, non-HCQ group had shorter median time to negative PCR, 19 days (IQR:8) when compared to HCQ sub-groups mild and severe, (*p* 0.004 and 0.002) respectively). Univariate analysis of “time to negative PCR” shows that it is significantly correlated with older age, having co-morbidities, cough, antibiotic use, higher CRP, and D-dimer (all *p*<0.05). In the multivariate regression model (F=6.8, *P*<0.002, R^2^=0.20), only two variables remain significant: HCQ and age. Being in HCQ group would delay the time to negative PCR by 7 days (95%CI: 2-12) and with every year increase in the age, the time to negative PCR would be delayed by 0.12 (95%CI: 0.017-0.22).

## Discussion

Since the start of COVID-19 pandemic, there has been an explosion of published research on different aspect of SARS-CoV2 infection. After the initial optimism of Hydroxychloroquine as an effective treatment for COVID-19, the subsequent studies have been conflicting. The negative studies and concern for harmful effects led to exclusion of Hydroxychloroquine arm of multinational SOLIDARITY trial and FDA revoked the emergency use authorization (EUA) for Hydroxychloroquine and Chloroquine for treating certain hospitalized patients with COVID-19 when a clinical trial was unavailable or participation in a clinical trial was not feasible. A recent large observational multihospital study from USA shows reduction in COVID-19 associated mortality with the use of Hydroxychloroquine alone or in combination with Azithromycin.^11^ The effect of Hydroxychloroquine and Azithromycin on duration from symptom onset to negative SARS-CoV-2 RT-PCR is variable. While some small studies have shown faster viral clearance with the use of Hydroxychloroquine, the others suggest no effect or delay in viral clearance. An observational French study suggests the reduced viral load with use of combination of Hydroxychloroquine and Azithromycin in mild COVID-19 disease.^12^ Another study with historical control shows that chloroquine shortens the time to viral clearance.^13^ A small randomized trial comparing Chloroquine and lopinavir/ritonavir suggests faster viral clearance with Chloroquine.^14^ Other studies including two randomized control trials did not show significant effect on time to viral RNA clearance.^15-18^ A small retrospective study suggests delayed viral clearance with the use of Hydroxychloroquine.^19^

The results of this study suggest that Hydroxychloroquine in combination with Azithromycin may delay the viral clearance and prolong the duration from symptom onset to negative PCR as compared to patients who are not given Hydroxychloroquine. The fact that non-HCQ group had shorter duration to negative PCR when compared to HCQ group suggests that Hydroxychloroquine and Azithromycin combination may have a role in delaying the virus clearance. Age may have effect on delaying viral clearance as older patients ≥60 year had longer duration to viral clearance compared to patients <60 year in HCQ group. In the multivariate regression model (F=6.8, *P*<0.002, R^2^=0.20), only two variables were significant: HCQ and age. Being in HCQ group would delay the time to negative PCR by 7 days (95%CI: 2-12) and with every year increase in the age, the time to PCR negative would be delayed by 0.12 (95%CI: 0.017-0.22). There is a possibility that Hydroxychloroquine and/or Azithromycin may delay the virus clearance through its immunomodulatory effect especially a specific immune suppression necessary to combat SARS-CoV-2 virus.^20^

## Conclusion

Hydroxychloroquine and Azithromycin may have a role in delaying SARS-CoV-2 virus clearance, prolonging the time from symptom onset to negative PCR in patients hospitalized with COVID-19. Larger studies are needed to investigate this specific aspect of COVID-19 patients treated with these commonly used therapeutic agents.

## Data Availability

Data is available for review

## Limitation of study

The main limitation of the study is the small sample size. Selection bias and confounding factors may have played a role. There is a possibility of subjective bias in obtaining the history of exact date of symptom onset.

